# Methylome analysis for prediction of long and short-term survival in glioblastoma patients from the Nordic trial

**DOI:** 10.1101/2022.02.21.22271286

**Authors:** Małgorzata Łysiak, Jyotirmoy Das, Annika Malmström, Peter Söderkvist

**Affiliations:** Department of Biomedical and Clinical Sciences, Linköping University, Linköping, Sweden; Bioinformatics Unit (Core Facility), Linköping University, Linköping, Sweden; Clinical Genomics Linköping, SciLife Laboratory, Department of Biomedical and Clinical Sciences, Linköping University, Linköping, Sweden; Department of Advanced Home Care, Linköping University, Linköping, Sweden

**Keywords:** glioblastoma, methylation, long-term survival, short-term survival, temozolomide, radiotherapy

## Abstract

Patients with glioblastoma (GBM) have a short survival, but even among patients receiving the same therapies and with good prognostic factors, one can find those with exceptionally short and long survival.

In the Nordic trial, patients with GBM, 60 years or older, were randomized between 2 radiotherapy arms or TMZ. We selected 59 patients, equally distributed between the 3 treatment arms and MGMT promoter methylation status, with good prognostic factors, but with short or long survival. We performed methylation profiling with the Illumina Infinium Methylation EPIC BeadChip arrays in conjunction with a methylation-based CNS tumor classifier, analysis of differentially methylated CpG sites (DMCs) and pathway enrichment analysis.

Samples classified as non-GBM IDH wildtype were excluded and in the analysis of long vs. short survivors with documented progression or tumor-related death, we found DMCs in the TMZ, MGMT promoter methylated group (123,510), as well as in the 60Gy, MGMT promoter unmethylated group (4,086) and 34Gy, MGMT promoter methylated group (39,649). The joint analysis of the RT arms revealed 319 DMCs in the MGMT unmethylated group but no differences for MGMT promoter methylated samples, or in any of the analyses independent of MGMT status. Interestingly, in the long-term survivors with methylated MGMT promoter treated with TMZ we found hypermethylation of the Wnt signaling and the platelet activation, signaling and aggregation pathways.

We identified DMCs for both TMZ and RT treated patients. Further systematic analysis of larger patient cohorts is necessary for confirmation of their predictive properties.

## Introduction

Glioblastoma (GBM) remains the most common and deadliest among gliomas. The peak incidence is in the individuals above 65 years old and only about 5.6% of patients reach 5-year survival [1]. Unfortunately, the treatment options are also limited and so far, efforts have not provided satisfactory survival benefits, leaving the median survival at 1.2 years [2,3]. Standard therapy for GBM patients consists of gross total resection, if feasible, followed by concomitant radiotherapy (RT) and chemotherapy with the alkylating agent temozolomide (TMZ) and additional six cycles of TMZ [4]. For fit patients, additional treatment with tumor treating fields is an option, further prolonging survival for the selected group by near 5 months [5]. Choosing the best possible treatment is crucial, especially for elderly patients often burdened with comorbidities, where the combined treatment is not expected to be tolerated. The known positive prognostic factors include the extent of surgery, age, performance status and sex of the patient [2]. Thus far, the only predictive biomarker associated with response to TMZ treatment is the methylation status of the *O*^*6*^*-methylguanine-DNA-methyltransferase* (*MGMT*) gene and methylated MGMT (m-MGMT) is associated with better overall survival (OS) [6,7], although varying responses among patients are still observed.

Notably, the importance of epigenetic changes in gliomas has been emphasized by recent discoveries [8-11]. The current classification of brain tumors incorporates molecular biomarkers, i.e., mutations of isocitrate dehydrogenase 1 or 2 (IDH1/2) and 1p/19q codeletion [12], and methylome profiling adds a promising alternative, which introduces refinement and subclassification to the present classification system [8]. Another example is glioma cytosine-phosphate-guanine (CpG) island methylator phenotype (G-CIMP), which entails genome-wide hypermethylation of the CpG islands, especially common among IDH1/2 mutated gliomas and associated with better outcome [13-15]. There are also other indications that methylome profiling could aid in selection of patients with better prognosis within the same diagnostic entity, e.g., by using the methylation differences found in short- and long-term survivors (STS and LTS, respectively) with GBM [16-18]. Moreover, changes in the methylation profiles between primary and recurrent GBM have been reported, with common occurrence of switches between methylation subclasses found at progression [18].

Age is one of the prognostic factors and younger GBM patients are characterized by better outcomes [1,2]. Aging is also reflected in the methylation state of the genome, so called epigenetic age [19]. Methylation of specific CpG sites undergoes age-dependent changes, which can be quantified and expressed through the epigenetic age [20]. Acceleration of the epigenetic age, which is the difference between the epigenetic age and chronological age, has been reported in many diseases, such as Alzheimer’s disease [21] or Parkinson’s disease [22]. It has also been shown to associate with cancer, mortality in cardiovascular diseases [23,24], and patients’ outcome in gliomas [25,26].

In the Nordic trial, patients 60 years or older diagnosed with GBM were randomized between two different dose regimens of RT or to TMZ treatment alone. Constituting a unique cohort, we decided to analyze the global methylation status of tumors from LTS and STS using Illumina EPIC bead arrays, to identify potential methylation-based biomarkers or profiles related to treatment and outcome.

## Materials and methods

### Patients

All patients included in this study were participants of the Nordic randomized, phase 3 trial registered under the number ISRCTN81470623 [6], which compared 3 treatment modalities for newly diagnosed GBM patients with age 60 and above (TMZ vs. standard RT 60Gy vs. hypofractionated RT 34Gy). We selected 59 patients with available formalin fixed paraffin embedded (FFPE) tumor tissue from the 3 treatment arms: 20 patients from the TMZ and 34Gy arms and 19 patients from the 60Gy arm (Figure 1). All patients underwent tumor resection and had WHO performance status 0-1, both considered as good prognostic factors. All patients were IDH1 negative, tested by immunohistochemistry [6]. Patients were selected, so that for each treatment arm half of them belonged to the long survival group and half to the short survival group, and in each survival group there would be an equal number of patients with methylated MGMT promoter (m-MGMT) and unmethylated MGMT promoter (u-MGMT) (Table 1). The MGMT methylation status was analyzed with the MDxHealth method, Liège, Belgium, as mentioned in [6].

**Table 1.** Patient characteristics.

| Treatment | Survival group | MGMT status | N= | Males | Mean age at diagnosis (range) [years] | Median survival (range) [months] |
| --- | --- | --- | --- | --- | --- | --- |
| TMZ | LTS | Methylated | 5 | 3 | 64 (60-69) | 20.1 (13.7-125.9) |
|  |  | Unmethylated | 5 | 3 | 72 (67-77) | 13.3 (9.9-18.9) |
|  | STS | Methylated | 5 | 2 | 70.6 (67-78) | 9.2 (3.5-10.4) |
|  |  | Unmethylated | 5 | 3 | 66.8 (64-69) | 2.9 (2.1-3.8) |
| 34Gy | LTS | Methylated | 5 | 1 | 69.2 (63-77) | 14.6 (12.6-24.3) |
|  |  | Unmethylated | 5 | 3 | 72.4 (69-81) | 14.6 (12.6-35.7) |
|  | STS | Methylated | 5 | 2 | 72.4 (65-77) | 4 (1.3-5.5) |
|  |  | Unmethylated | 5 | 2 | 68.6 (64-74) | 4.6 (3.3-5.1) |
| 60Gy | LTS | Methylated | 5 | 2 | 64.2 (60-72) | 14 (12.1-16.6) |
|  |  | Unmethylated | 5 | 4 | 64.6 (60-70) | 17.9 (15.6-33.4) |
|  | STS | Methylated | 4 | 3 | 69.3 (62-73) | 7.8 (6.6-10.1) |
|  |  | Unmethylated | 5 | 4 | 69.2 (65-74) | 1.7 (1.1-4.2) |

**Figure 1.**
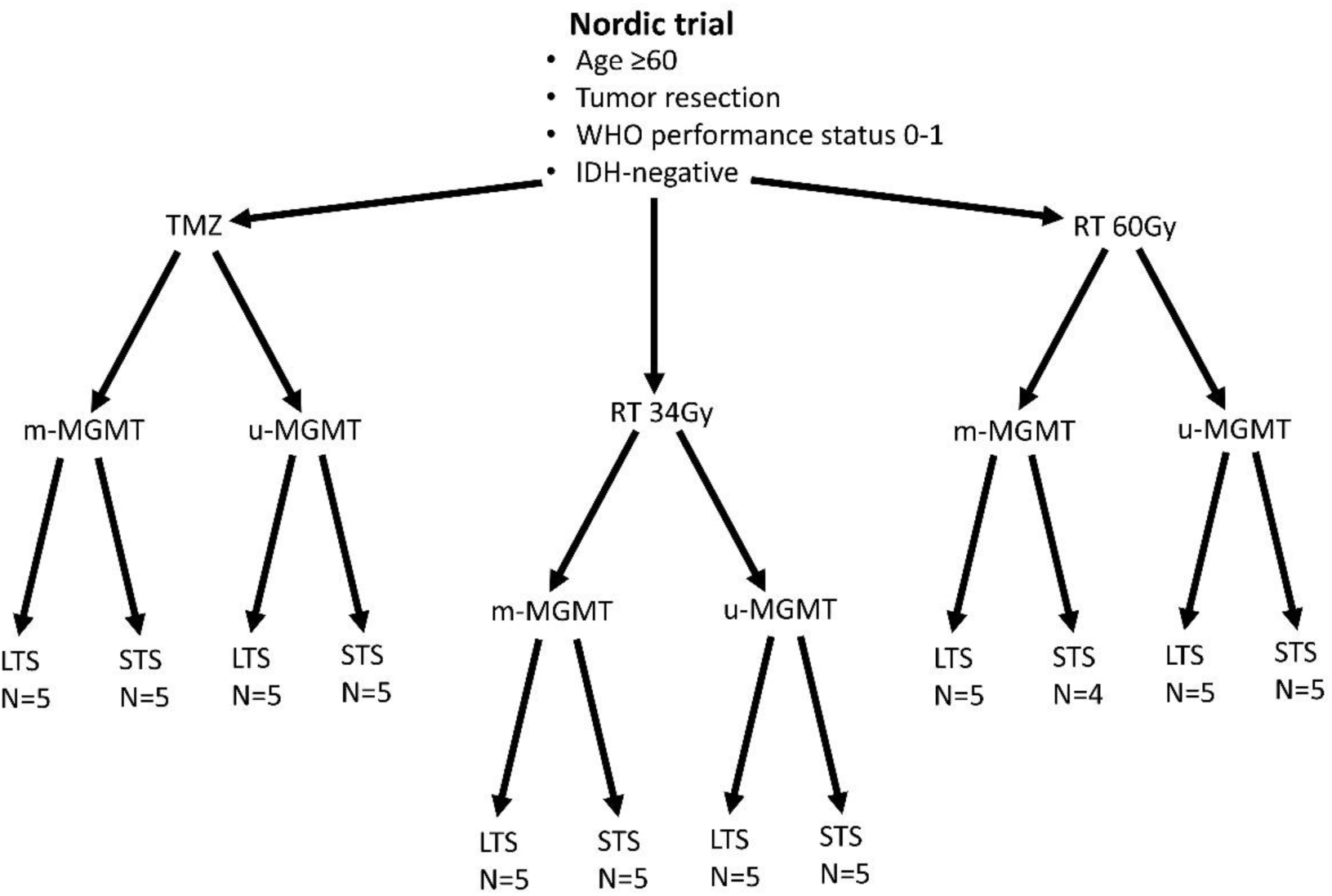
Patients from the Nordic trial included in the study. Patients with good prognostic factors were selected from each treatment arm (temozolomide -TMZ, radiotherapy 34Gy-RT 34Gy and radiotherapy 60Gy-RT60Gy). Half of the tumors had methylated MGMT promoter (m-MGMT) and half had unmethylated MGMT (u-MGMT). These patients were further divided into long-term survivors (LTS) and short-term survivors (STS). N-number of patients

### DNA methylation analysis

DNA was extracted from 2 tumor tissue sections of 10μm each, using the Maxwell FFPE DNA Purification Kit (Promega) according to the manufacturer’s protocol but with a double amount of proteinase K (40mg/ml). DNA quantity and quality were checked with NanoDrop ND-1000 Spectrophotometer (ThermoFisher) and Quantus Fluorometer (Promega), as well as with the Infinium FFPE QC Kit (Illumina). A total of 250-500ng of DNA was subjected to bisulfite conversion using the EZ DNA methylation kit (Zymo Research) and genome wide DNA methylation was assessed with the Infinium MethylationEPIC BeadChip Kit (Illumina) complemented by the Infinium HD FFPE DNA Restore Kit as per manufacturer’s protocol. The BeadChip arrays were scanned on the NextSeq 550 (Illumina) and DNA methylation data in the form of IDAT files (intensity data files that contain green and red signals from methylated and unmethylated CpG sites) were uploaded to the online classifier v11b4 [8], where MGMT methylation status was also assessed, with the MGMT-STP27 algorithm [27].

### Differential methylation analysis

The IDAT files from Illumina HumanMethylation EPIC arrays were also analyzed using R (v4.0.3) [28] and Bioconductor packages (v3.14) [29], e.g., Chip Analysis Methylation Pipeline (ChAMP) analysis package (v2.19.3) [30]. The files were pre-processed in ChAMP to filter out CpGs with detection p-value >0.01, as well as SNP CpGs, unbound and multi-hit CpGs and all CpGs from sex chromosomes. After filtration, the quality check was performed, and the files were normalized with the beta-mixture quantile normalization (BMIQ) function. The β- and M-values of the samples were calculated against each CpG per sample. Batch effects were corrected for with the *runCombat* function. The differential methylation analysis was calculated with the linear modeling (*limFit*) and *eBayes* algorithm, comparing two groups from the phenotypic dataset and singular value decomposition (SVD) analysis was performed to check for confounders (e.g., age, sex) (Supplementary Data, Figure S1). The differential methylation analysis was performed first for all samples that were classified as GBM, IDH wild-type and then, after removal of patients that died due to causes other than tumor, for 3 samples from each group with the extreme survival, that is in LTS the 3 with the longest survival and STS, the 3 with the shortest survival. The latter analysis found differentially methylated CpGs (DMCs) in the comparison of LTS vs. STS within each treatment arm with methylated or unmethylated MGMT promoter, hence further analysis was based on these. The DMCs were considered significant at the Bonferroni-Hochberg corrected *p*-value (*p*-value_BH_) <0.05. The hierarchical cluster analysis was performed using the Euclidean distance within the *ape* package (v5.0) [31] in R.

### Structural annotation

We used *AnnotationDbi* package (v1.54.1) [32] to annotate DMCs and in-house script to visualize their genomic distribution. The statistically significant DMCs (*p*-value_BH_ < 0.05) were used to create the volcano plot with the mean methylation difference (Δmmd) ≥|0.3|) using the *EnhancedVolcano* package (v1.10.0) [33] in R. The cut-off of Δmmd was calculated using the β-value distribution of all samples with the mean ±2SD (Supplementary Data, Figure S2 and Supplementary Data, Table S1).

The R package *ComplexHeatmap* (v2.8.0) [34] was used to create the heatmap from individual β-values of DMCs (*p*-value_BH_ < 0.05; (Δmmd) ≥|0.3|).

### Pathway enrichment and correlation analysis

To reduce the number of DMCs, we first filtered out DMCs based on the genomic location, leaving only those from the 5’-untranslated region (5’-UTR) and transcription start site (TSS) regions (TSS200 and TSS1500). Further, we applied the Δmmd cut-off score (as described above). The DMCs were converted to their respective official gene symbols (hereafter called DMGs, differentially methylated genes) and the list (without/with Δmmd values) was used for the pathway enrichment analysis. The Reactome database (v78) [35] was applied to perform the gene set enrichment analysis using the c*lusterProfiler* package (v4.0.5) [36] in R (v4.1) with the default parameters setup (e.g., 1000 permutations and *p*-value_BH_ < 0.05). Results were visualized using ggplot2 (v3.3.3) [37] in-house script.

### Epigenetic age calculation

The epigenetic age was calculated for the same set of samples that were used in the differential methylation analysis (3 samples from STS and LTS per group). We used methylation data obtained in this study and followed the methods published for three epigenetic clocks, namely Horvath [20], Hannum [39] and PhenoAge [40]. The epigenetic age acceleration was calculated as the difference between the epigenetic age and chronological age, given in years. Mean chronological and epigenetic ages were compared with the two-tailed Student-t test and *p-*value<0.05 were considered significant. Calculations were performed with IBM SPSS (v.26). Ethical approval for the Nordic trial and molecular analyses were previously obtained (99086, M11-06 T40-09 and 2011/32-32).

## Results

### Methylation-based classification

Histological review in the primary analysis of the trial material classified 58 out of 59 samples included in this study as GBM grade 4 and one sample was classified as astrocytoma grade 3 but all samples were IDH1 mutation negative. Methylation data for all samples passed the quality control and were uploaded to the brain tumor classifier (https://www.molecularneuropathology.org/mnp) [8]. Methylation based classification placed most of the samples in the GBM, IDH wildtype class, including the mentioned astrocytoma.

One sample was classified as anaplastic pilocytic astrocytoma and confirmative sequencing of IDH1 and IDH2 displayed the absence of mutation in accordance with the immunohistochemistry result. In three cases, samples were classified as “control tissue” probably due to low tumor cell content and were excluded. The analysis of methylation subclasses revealed that 21 samples belonged to only one subclass, with 2 being midline GBM. The remaining (n=34) GBM IDH wildtype samples had 2 or 3 subclasses assigned to them. The MGMT methylation analysis results were concordant between the MDxHealth method and MGMT-STP27 for all but one sample, even for the samples that were classified as control tissue.

### Differential methylation analysis

We decided to only include samples, where progressive diseases and/or death caused by the tumor had been reported because, especially in the STS groups, the true relationship between the tumor’s methylation profile and survival could be compromised. After removing cases, where death was caused by co-morbidity or complications (e.g., infection, pulmonary embolism), we decided to include only three samples with extreme survival times from each treatment arm and MGMT group, as survival times, especially in the RT arms, had relatively small spread. and

We compared LTS and STS samples in the separated treatment arms (TMZ, 34Gy, 60Gy, and combined RT) and within each treatment we compared m-MGMT and u-MGMT samples separately and in combination. DMCs between LTS and STS were identified in the TMZ arm, m-MGMT; 34Gy, m-MGMT; 60Gy, u-MGMT and in the combined RT arm, u-MGMT. The highest number of DMCs were found in the TMZ group (Supplementary Data, Table S1). The cofactor analysis showed that the differential methylation analysis was not influenced by included confounders (age, sex, death by the tumor/progressive disease) (Supplementary Data, Figure S1). Upon the structural annotation of DMCs, we found that they were similarly distributed throughout the genome (Figure 2), with the majority of DMCs found in the gene bodies and intergenic regions. Approximately 10% of DMCs were found in the TSS1500 and up to 8% in the 5’UTR, with both regions being of regulatory importance for gene expression due to the location of gene promoters. We reduced the number of DMCs for further analysis by filtering them out based on the genomic location (TSS1500, TSS 200 and 5’-UTR) and Δmmd cut-off values. Inspection of density plots created from filtered data revealed that a majority of DMCs were hypermethylated in LTS in the TMZ and 34Gy, m-MGMT groups and hypomethylated in the LTS from 60Gy, u-MGMT group. All DMCs were hypomethylated in the comparison of LTS vs. STS in the combined RT group with u-MGMT (Supplementary Data, Figure S2). Next, we performed hierarchical cluster analysis on filtered DMCs (Figure 3). We observed that in the TMZ, m-MGMT group, data formed 4 clusters of DMCs between LTS and STS. We also found that for the joint RT group with u-MGMT, clusters separated LTS and STS but not the radiation doses (34Gy and 60Gy), emphasizing the importance of the treatment modality itself.

**Figure 2.**
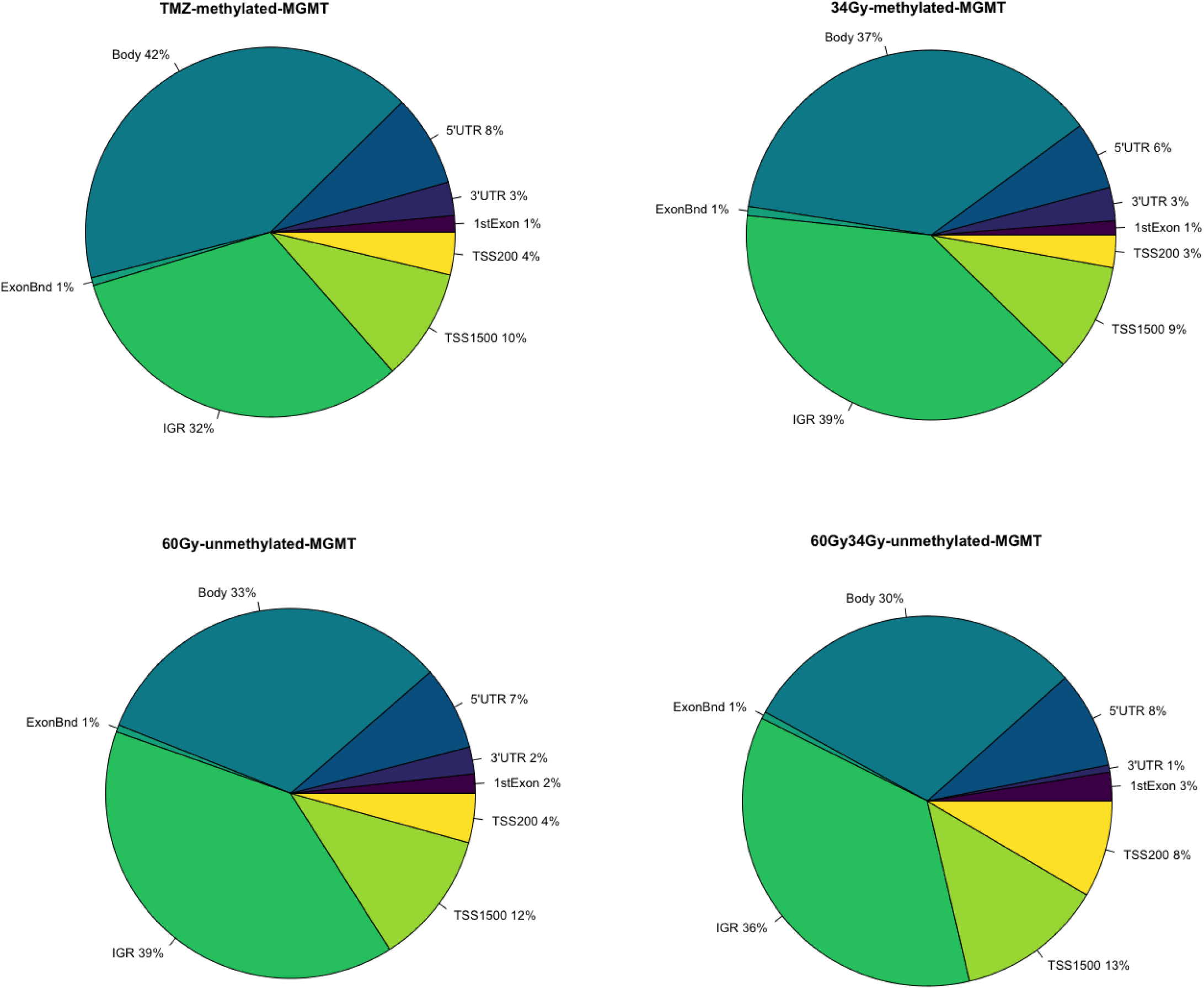
Pie charts representing the structural genomic distribution of DMCs discovered in samples with exceptionally long (n=3) and short (n=3) survival within different treatment arms and with specified MGMT promoter methylation status. TSS200-200 bases upstream transcription start site, TSS1500-1500 bases upstream transcription start site, UTR-untranslated region, IGR-intergenic region

**Figure 3.**
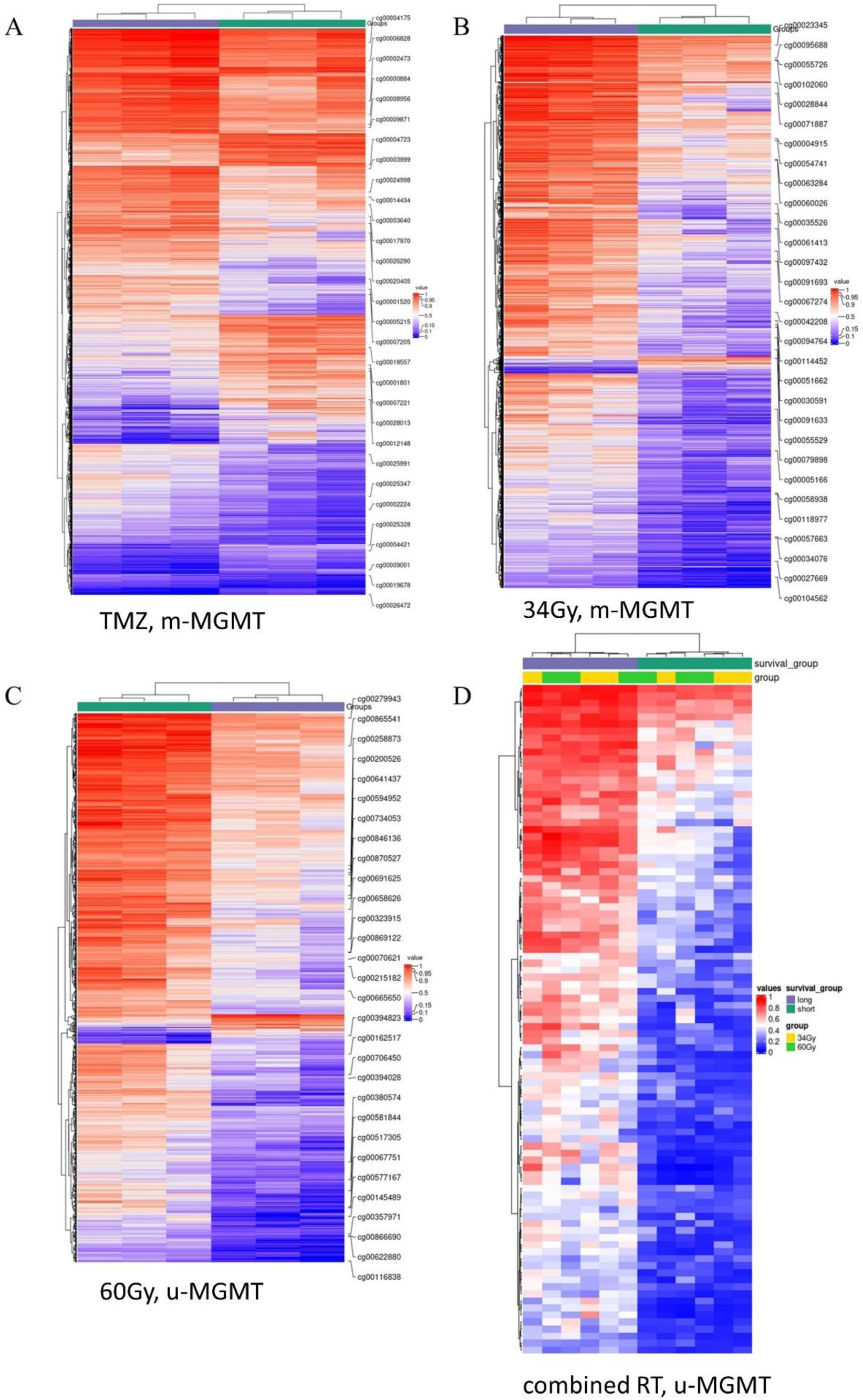
Heatmaps representing β-values of the DMCs for (A) TMZ, m-MGMT, (B) 34Gy, m-MGMT, (C) 60Gy, u-MGMT, (D) RT, u-MGMT.

### Pathway enrichment analysis

Next, we used the Reactome database to investigate pathway enrichment among filtered DMGs, which were obtained from the annotated DMCs. There were four pathways enriched in the hypermethylated DMGs among LTS from the TMZ, m-MGMT group, namely metabolism; platelet activation, signaling and aggregation; signaling by WNT and signal transduction. In the same group analysis (LTS from the TMZ, m-MGMT), we found 37 pathways enriched in the hypomethylated DMGs, e.g., immune system and Rho signaling pathways (Supplementary Data, Table S2). In the hypermethylated DMGs in LTS in the treatment of 34Gy with m-MGMT we found two enriched pathways (immune system and class B/2 secretin family receptors) (Supplementary Data, Table S2). We did not find any enriched pathways for the remaining groups. Due to stringent filters applied initially to DMCs and low number of detected enriched pathways, we decided to include DMCs removed by the Δmmd cut-off and repeat the analysis. Consequently, the number of enriched pathways increased, both for the TMZ, m-MGMT and 34Gy, m-MGMT groups (Figure 4), but no pathways were enriched in the 60Gy and combined RT groups. We compared the lists of enriched pathways and found seven that were common for the TMZ and 34Gy (m-MGMT) arms, with 3 of them involved in G-protein coupled receptors (GPCR) signaling (Figure 4). One of the hypermethylated pathways found to be enriched in the LTS from the TMZ group with m-MGMT was WNT signalling. We wanted to check whether there was a correlation between methylation of DMGs from the enrichment core and the expression of these, but gene expression data were not available for our samples. Instead, we used 51 primary, IDH wild-type GBM with RNA-seq and 450k methylation array data from TCGA, accessed via the SMART App website [38]. First, we identified which of the DMCs from the enrichment core were also covered by 450k bead chip arrays, since our results were based on the newer design, 850k methylation array. The overlapping DMCs were found for 9 genes, which were then analysed with the Spearman correlation coefficient to find if the promoter methylation status of the genes correlated with the corresponding mRNA expression, at a significance level p<0.05. A negative correlation was found only for the WNT2 gene (R=-0.37, p=0.0067).

**Figure 4.**
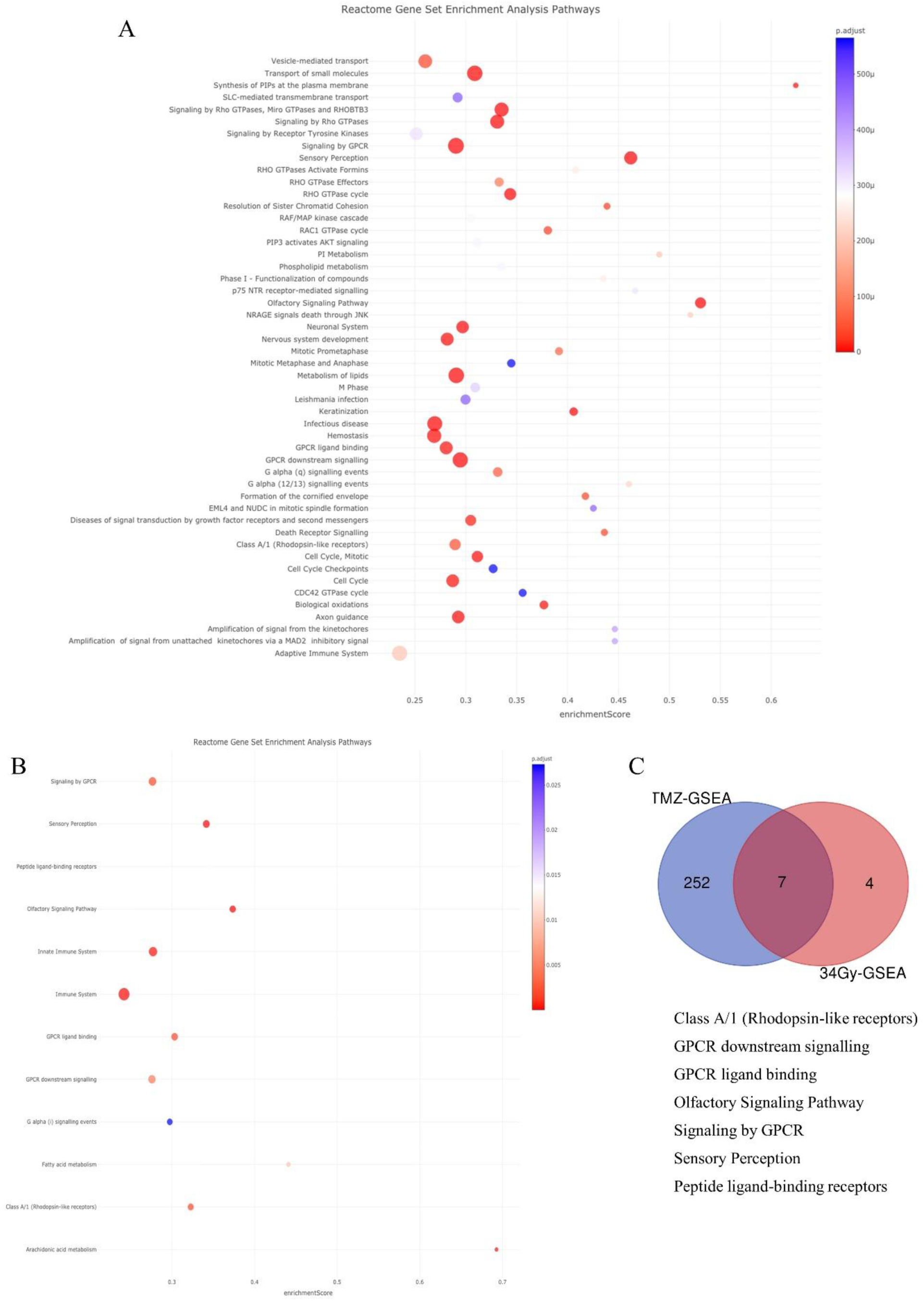
Results of pathway enrichment analysis for DMGs from (A) TMZ, m-MGMT (top 50 pathways) and (B) 34Gy, m-MGMT (B). Venn diagram (C) shows the number of unique and shared enriched pathways between the two groups (TMZ (blue) and 34Gy (red)) and lists the latter.

### Epigenetic age

The cofactor analysis showed that chronological age did not affect DMCs (Supplementary Data, Figure S1), however, molecular alterations in cancer cells may affect the epigenetic age. This prompted us to analyze the epigenetic age of the groups consisting of 3 tumor samples, which involved three different epigenetic clocks, Horvath [20], Hannum [39] and PhenoAge [40]. The calculated epigenetic ages were compared with the chronological age of the patient. According to the Horvath algorithm, all samples had a higher epigenetic than chronological age (Figure 5) but there was no difference between epigenetic age in LTS and STS. There were also no differences in the age acceleration (Supplementary Data, Table S3). In the results from the Hannum and PhenoAge epigenetic clocks, both, epigenetic age acceleration and deceleration (epigenetic age<chronological age) were observed, with STS groups usually characterized by lower mean epigenetic age (Figure 5 and Supplementary Data, Table S3). Significant differences were discovered only in the 34Gy, u-MGMT group (Hannum and PhenoAge) and in the combined RT, u-MGMT group (Hannum). In these cases, STS showed epigenetic age deceleration and lower mean epigenetic ages than LTS.

**Figure 5.**
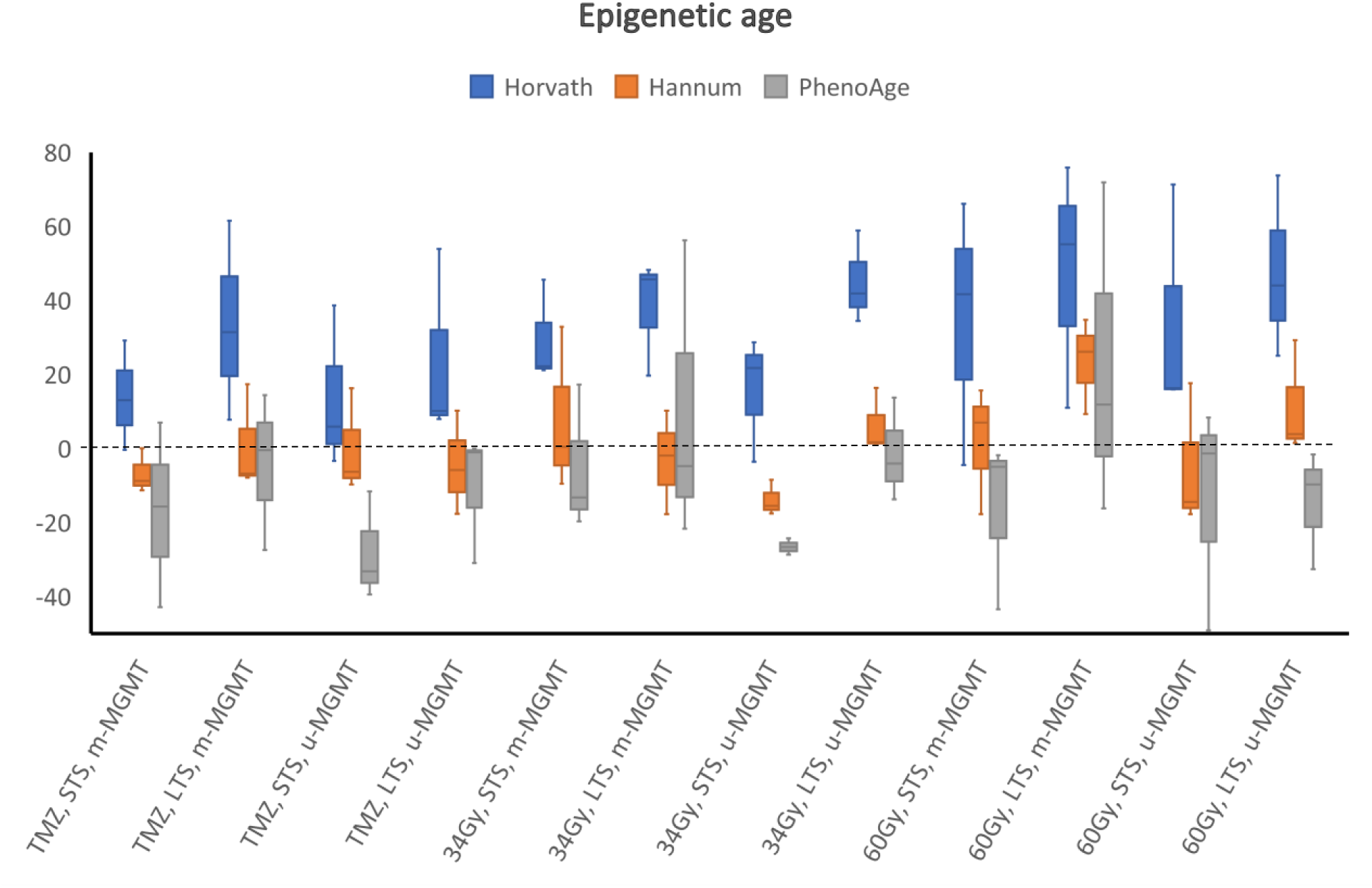
Results of the epigenetic age analyses. The epigenetic age acceleration (difference between the epigenetic age and the chronological age) is shown in years on the y-axis, horizontal lines represent the median values, boxplots represent the 1st and 3^rd^ quartile, errors are represented by the T-bars. All data above the dashed line represents an acceleration in epigenetic age, and data below the dotted line represents a deceleration in epigenetic age.

## Discussion

In recent years, an increase in the use of methylome profiling has been observed. The widely known brain tumor classifier proposed by Capper et al. [8] allows for a more precise classification of brain malignancies. Undoubtedly, methylation of the MGMT promoter remains the most important biomarker for GBM, which predicts effect of TMZ treatment [2,6,7]. However, we lack validated predictive biomarkers for patients with u-MGMT tumors or treated with RT. Here, we employed the Capper classifier [8], and differential methylation to identify biomarkers predisposing to good treatment response or indicating resistance to therapy.

Inter- and intratumor heterogeneity of GBM plays an important role in treatment resistance and relapse of the disease. A previous study on spatially separated biopsies showed that methylation heterogeneity is common in GBM, though it seems to affect only the subclassification, whilst all samples remain classified as GBM IDH wildtype and MGMT methylation status also remains stable [42]. This is in line with our results from the methylation-based classification, which showed that in 34 samples more than one subclass was identified. Interestingly, it has recently been reported that treatments such as chemo- or radiotherapy, and even hypoxia, as sources of stress for cancer cells, may induce methylation changes that likely contribute to heterogeneity among tumor cells [43] and possibly patients’ outcome.

Our analysis revealed that the WNT signaling pathway was hypermethylated in LTS treated with TMZ and with m-MGMT, implicating silencing of WNT signaling and indicating the importance of a maintained WNT pathway activity in STS. This aligns with the previously presented results by Shinavi et al. [16], where hypermethylation of the promoter of the DKK2 gene, an antagonist of the WNT pathway, was found in STS. In TCGA data, we found a negative correlation between methylation of the sites corresponding to hypermethylated CpGs in our samples and the expression of the WNT2 gene. In mice, WNT2 drives the proliferation of progenitor cells and development of midbrain, possibly having similar effects in humans [44]. WNT signaling is crucial during embryonal development and is commonly altered in cancers [45]. In GBM, activation of the WNT pathway is necessary for the maintenance of glioblastoma cancer stem cells, which greatly contribute to treatment resistance [46]. Importantly, methylation may be the main regulatory mechanism of the WNT pathway in GBM, as mutations are rare [46]. GBM is also proposed to originate from the subventricular zone (SVZ), an area supporting neural progenitor cells [47], and tumor proximity to the SVZ has been linked to STS [48]. Methylation of gene promoters in newly diagnosed GBM and their impact on gene expression, and survival of patients treated with concomitant radiochemotherapy, was also previously reported by Etcheverry et al. [9]. They found six CpG sites, for which methylation was associated with decreased survival, and two of the CpG sites were localized in the SOX10 gene promoter. We did not find SOX10 among DMGs in any of the treatment arms but active WNT signaling acts inhibitory on SOX10 expression [49]. To further emphasize this signaling pathway, study by Wu et al. [11] showed, that SOX10 acts as a master regulator of the receptor tyrosine kinase I (RTK1) subtype of GBM, which often harbors platelet derived growth factor receptor alpha amplification [11].

Another interesting finding is hypermethylation of the platelet activation, signaling and aggregation pathway in LTS with m-MGMT promoter treated with TMZ. Up to 30% of GBM patients experience venous thromboembolism, making it one of the most common and serious complications, which may influence survival [50]. Hypermethylation of the platelet activation pathway may therefore have a protective function.

Among the hypomethylated enriched pathways we found some that were labelled as neuronal system, neurotransmitter receptors, transmission across chemical synapses and potassium channels, all of which have been shown to partake in GBM development and progression [51,52]. In our study however, these pathways associated to LTS. It should be noted that in most of the published studies mentioned here, STS is referred to patients with a survival <1 year and LTS to those with a survival >3 years. In our study of elderly patients with GBM, median survival was 12 months and the survival range of 4.5-126 months for the most favorable group, those treated with TMZ and m-MGMT.

TMZ is most commonly used in GBM treatment due to its good blood brain barrier penetration and generally mild toxicity [53] and MGMT promoter methylation status is predictive for the treatment response to TMZ [6]. However, not all GBMs are tested for MGMT [54]. Chai et al. investigated the potential benefits of TMZ in patients with u-MGMT and showed that by using a methylation signature of 31 genes, it is possible to select patients with u-MGMT to obtain a survival similar to that of patients with m-MGMT [10]. Unfortunately, we did not find any DMCs separating LTS and STS in the TMZ arm with u-MGMT.

The pathway enriched in the hypermethylated DMGs in LTS from the 34Gy treatment arm with m-MGMT promoter, is the pathway of the secretin family receptors, a subgroup of G-protein coupled receptors (GPCR). Pathways involving GPCR are enriched in the TMZ and 34Gy arms when filtering conditions are less stringent. GPCR is the largest family of membrane proteins involved in cell metabolism, migration, neurotransmission, immune response, and cell differentiation [55]. Hypermethylation of these pathways may lead to downregulation of gene expression and decreased activity, potentially having a protective effect on the patients. In fact, GPCRs are explored in various studies as treatment targets for glioblastoma [55] but in our limited study, we did not find any DMCs in the combined RT group to indicate the importance of methylation of GPCR for RT outcome.

Although we did not find consistent and significant differences in the epigenetic age of LTS and STS in most of the analyzed treatment groups, we observed a general distortion in the epigenetic age in comparison to the chronological age. The magnitude and direction of epigenetic age changes was dependent on the applied algorithm but overall, we observed a trend towards lower epigenetic age in STS in comparison to LTS in all algorithms. The Horvath’s clock pioneered the field and can be universally used for different tissues [20], the Hannum’s clock is primarily designed for assessment of epigenetic age from blood and the PhenoAge model was built on phenotypic age, including many morbidities and mortality related factors. The Hannum’s method and PhenoAge both showed deceleration of epigenetic age instead of acceleration, which is likely dictated by the tissues and factors used for model development. However, the results from Horvath’s clock are in line with previous studies performed on gliomas, indicating acceleration of epigenetic age in tumor tissue [25,26]. The surprising effect of epigenetic age deceleration or lower age in STS could speculatively be the result of stem-like cells involvement in GBM development, since stem cells are characterized by lower epigenetic age [20].

The value of methylome profiling for brain tumors has been largely shown through the methylation-based CNS classifier [8]. Alhough our analyses are limited due to the number of samples, they also highlight methylation differences that exist between LTS and STS with GBM, that might be clinically relevant. Also, epigenetic age assessment may potentially be a valuable tool to select patients with good prognosis. However, systematic analysis of larger cohorts of patients with LTS and STS is necessary and warranted for validation of our findings.

## Data Availability

All data produced in the present study are available upon reasonable request to the authors and online at http://www.bioinfo-zs.com/smartapp/

## Acknowledgments

The authors would like to thank patients and research colleagues that participated in the Nordic trial [6]. We would also like to acknowledge Clinical Genomics Linköping, Science for Life Laboratory, Sweden for providing assistance with analysis and FORSS, LiU Cancer, Swedish Cancer Foundation and Lions research fund for funding this research.

## Supplementary Data

**Supplementary Figure S1.**
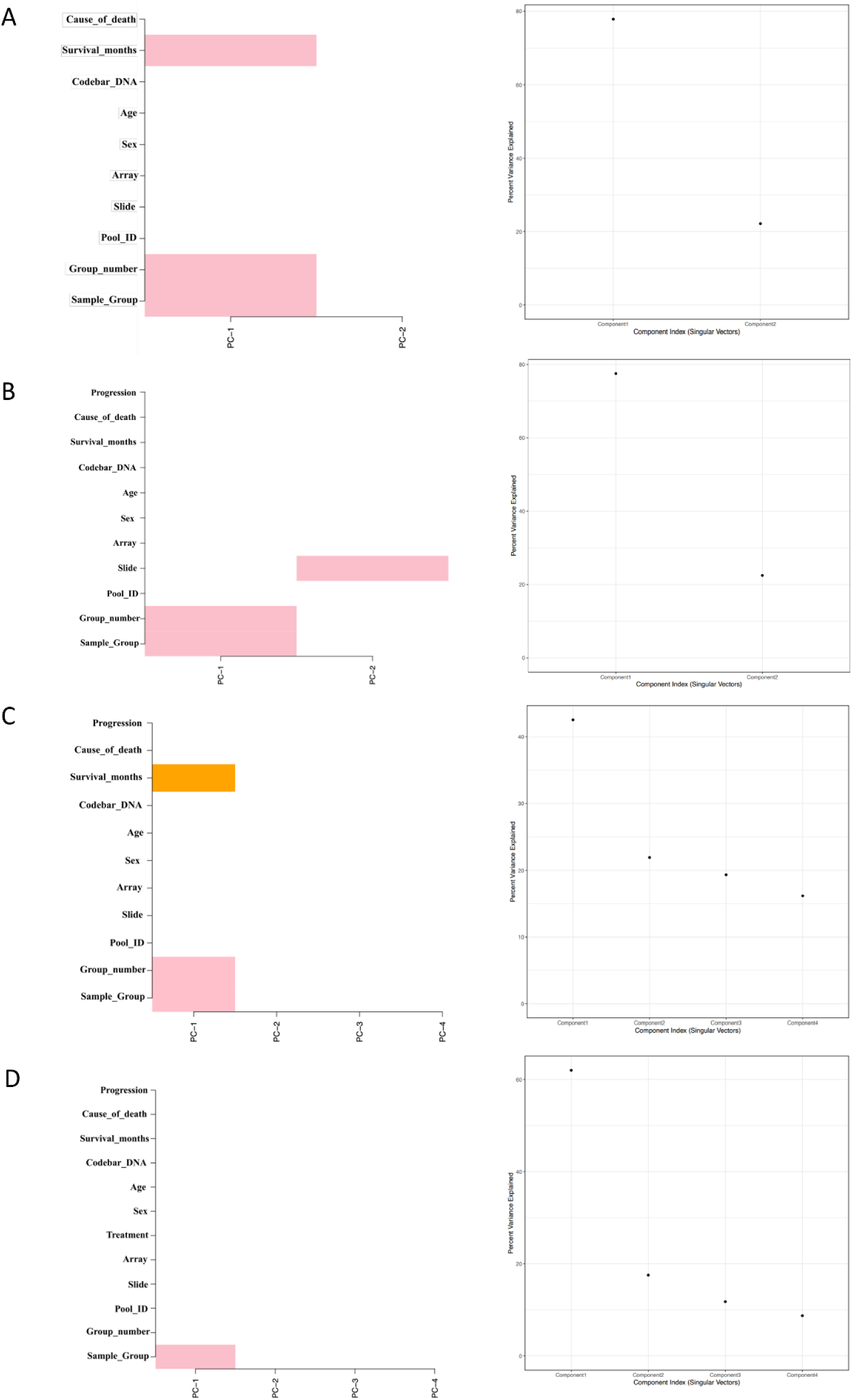
Results of the singular value decomposition (SVD) analysis checking for confounders in the groups with discovered DMCs; (A) TMZ, m-MGMT, (B) 34Gy, m-MGMT, (C) 60Gy, u-MGMT, (D) combined RT, u-MGMT.

**Supplementary Figure S2.**
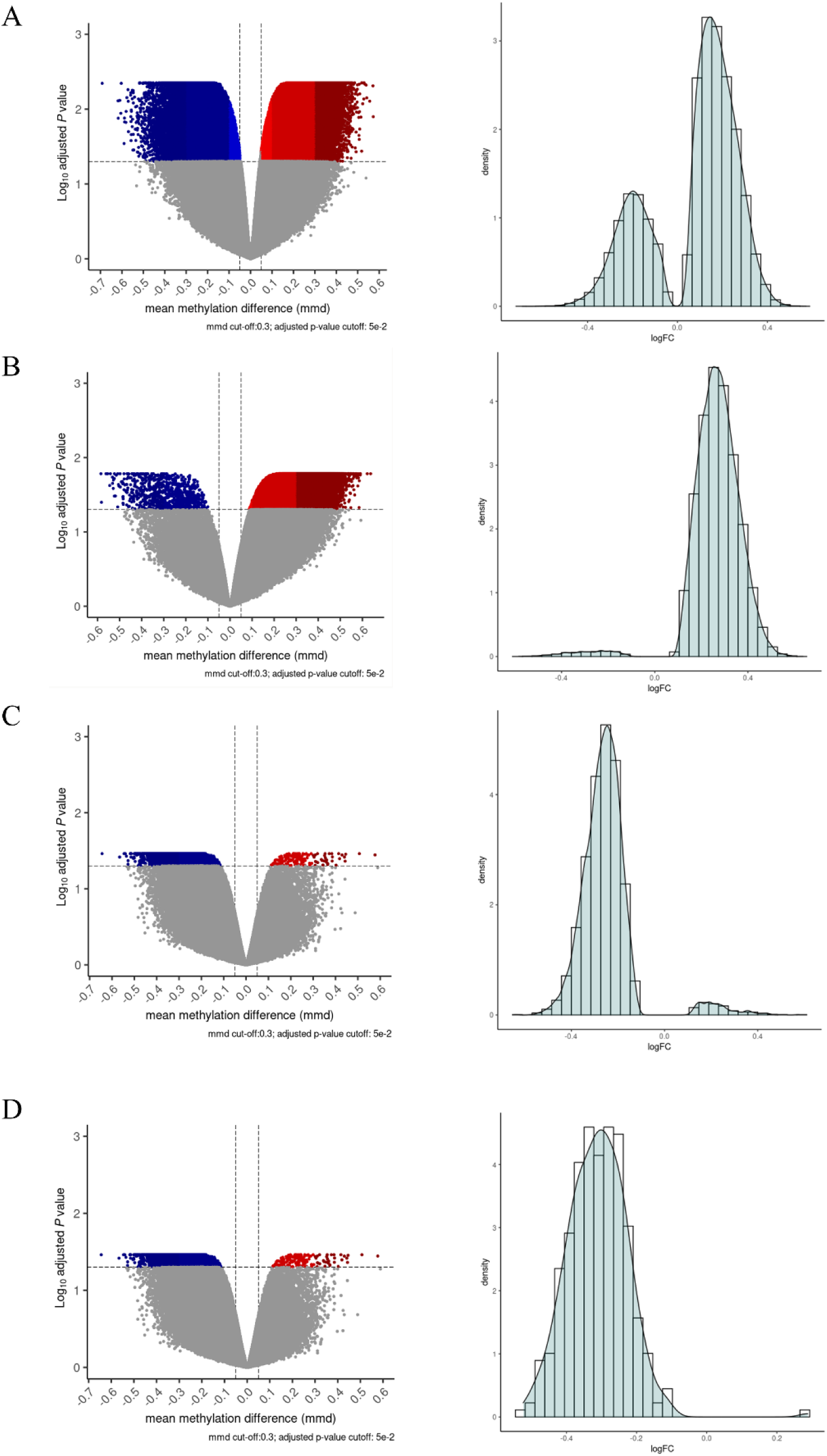
Volcano plots (left side) and density plots (right side) representing DMCs from (A) TMZ, m-MGMT, (B) 34Gy, m-MGMT, (C) 60Gy, u-MGMT, (D) RT, u-MGMT. Colored dots in the volcano plots in the darkest shades of blue and red represent DMCs fulfilling the cut-off requirements (*p*-value_BH_ < 0.05; (Δmmd) ≥|0.3|). Hypermethylated DMCs have positive values on the logFC axis in the density plots and hypomethylated DMCs have negative values logFC.

**Supplementary Table S1.** Number of differentially methylated CpGs (DMCs) and genes (DMGs) before and after location-based filtration.

| Comparison group<br>(LTS vs. STS) | Differentially methylated CpGs |  |  |  |
| --- | --- | --- | --- | --- |
|  | before filtering |  | only located at TSS and 5'UTR |  |
|  | DMCs | DMGs | DMCs | DMGs |
| TMZ, m-MGMT | 123510 | 18344 | 26626 | 11443 |
| 34Gy, m-MGMT | 39649 | 1104 | 39649 | 4915 |
| 60Gy, u-MGMT | 4086 | 1961 | 4086 | 816 |
| RT, u-MGMT | 319 | 181 | 319 | 82 |

**Supplementary Table S2.** Results of Reactome pathway enrichment analysis for filtered DMGs.

| Reactome ID | Reactome Pathway | Enrichment score | Adjusted $p$-value |
| --- | --- | --- | --- |
| **TMZ, m-MGMT, hypermethylated DMGs** |  |  |  |
| R-HSA-1430728 | Metabolism | 0.2310 | $4 \times 10^{-2}$ |
| R-HSA-76002 | Platelet activation, signaling and aggregation | 0.4427 | $4 \times 10^{-2}$ |
| R-HSA-162582 | Signal Transduction | 0.1964 | $4 \times 10^{-2}$ |
| R-HSA-195721 | Signaling by WNT | 0.5736 | $4 \times 10^{-2}$ |
| **TMZ, m-MGMT, hypomethylated DMGs** |  |  |  |
| R-HSA-168256 | Immune System | 0.2813 | $2 \times 10^{-7}$ |
| R-HSA-162582 | Signal Transduction | 0.2006 | $9 \times 10^{-6}$ |
| R-HSA-168249 | Innate Immune System | 0.2994 | $2 \times 10^{-4}$ |
| R-HSA-5653656 | Vesicle-mediated transport | 0.3626 | $2 \times 10^{-3}$ |
| R-HSA-597592 | Post-translational protein modification | 0.2861 | $2 \times 10^{-3}$ |
| R-HSA-1643685 | Disease | 0.2282 | $2 \times 10^{-3}$ |
| R-HSA-2682334 | EPH-Ephrin signaling | 0.6375 | $2 \times 10^{-3}$ |
| R-HSA-199991 | Membrane Trafficking | 0.3641 | $2 \times 10^{-3}$ |
| R-HSA-1280215 | Cytokine Signaling in Immune system | 0.2969 | $2 \times 10^{-3}$ |
| R-HSA-5663205 | Infectious disease | 0.2796 | $2 \times 10^{-3}$ |
| R-HSA-109582 | Hemostasis | 0.2902 | $2 \times 10^{-3}$ |
| R-HSA-392499 | Metabolism of proteins | 0.2262 | $3 \times 10^{-3}$ |
| R-HSA-6798695 | Neutrophil degranulation | 0.3438 | $3 \times 10^{-3}$ |
| R-HSA-112314 | Neurotransmitter receptors and postsynaptic signal transmission | 0.3745 | $3 \times 10^{-3}$ |
| R-HSA-112315 | Transmission across Chemical Synapses | 0.3340 | $3 \times 10^{-3}$ |
| R-HSA-1280218 | Adaptive Immune System | 0.2992 | $6 \times 10^{-3}$ |
| R-HSA-112316 | Neuronal System | 0.2673 | $6 \times 10^{-3}$ |
| R-HSA-74160 | Gene expression (Transcription) | 0.2246 | $6 \times 10^{-3}$ |
| R-HSA-9675108 | Nervous system development | 0.2727 | $9 \times 10^{-3}$ |
| R-HSA-212436 | Generic Transcription Pathway | 0.2258 | $9 \times 10^{-3}$ |
| R-HSA-73857 | RNA Polymerase II Transcription | 0.2258 | $9 \times 10^{-3}$ |
| R-HSA-1266738 | Developmental Biology | 0.2040 | $1 \times 10^{-2}$ |
| R-HSA-422475 | Axon guidance | 0.2704 | $1 \times 10^{-2}$ |
| R-HSA-202733 | Cell surface interactions at the vascular wall | 0.4847 | $2 \times 10^{-2}$ |
| R-HSA-9006934 | Signaling by Receptor Tyrosine Kinases | 0.2480 | $2 \times 10^{-2}$ |
| R-HSA-1296071 | Potassium Channels | 0.4801 | $2 \times 10^{-2}$ |
| R-HSA-977443 | GABA receptor activation | 0.4791 | $2 \times 10^{-2}$ |
| R-HSA-194315 | Signaling by Rho GTPases | 0.2796 | $2 \times 10^{-2}$ |
| R-HSA-9716542 | Signaling by Rho GTPases, Miro GTPases and RHOBTB3 | 0.2796 | $2 \times 10^{-2}$ |
| R-HSA-397014 | Muscle contraction | 0.3594 | $2 \times 10^{-2}$ |
| R-HSA-556833 | Metabolism of lipids | 0.3 | $2 \times 10^{-2}$ |
| R-HSA-449147 | Signaling by Interleukins | 0.3006 | $3 \times 10^{-2}$ |
| R-HSA-2029480 | Fcgamma receptor (FCGR) dependent phagocytosis | 0.4053 | $3 \times 10^{-2}$ |
| R-HSA-1430728 | Metabolism | 0.1621 | $3 \times 10^{-2}$ |
| R-HSA-1500931 | Cell-Cell communication | 0.4229 | $4 \times 10^{-2}$ |
| R-HSA-9658195 | Leishmania infection | 0.2667 | $4 \times 10^{-2}$ |
| R-HSA-418346 | Platelet homeostasis | 0.4401 | $4 \times 10^{-2}$ |
| **34Gy, m-MGMT, hypermethylated DMGs** |  |  |  |
| R-HSA-168256 | Immune System | 0.1954 | $4 \times 10^{-2}$ |
| R-HSA-373080 | Class B/2 (Secretin family receptors) | 0.6234 | $4 \times 10^{-2}$ |

**Supplementary Table S3.** Results of the epigenetic age calculations and comparisons of mean ages between LTS and STS.

| Treatment | MGMT status | Biological age [LTS vs. STS; p] | Horvath [LTS vs. STS; p] | Hannum [LTS vs. STS; p] | PhenoAge [LTS vs. STS; p] | Horvath acceleration [LTS vs. STS; p] | Hannum acceleration [LTS vs. STS; p] | PhenoAge acceleration [LTS vs. STS; p] |
| --- | --- | --- | --- | --- | --- | --- | --- | --- |
| TMZ | Methylated | 65 vs. 69; p=0.23 | 98.5 vs. 82.9; p=0.454 | 65.9 vs. 62.3; p=0.725 | 60.5 vs. 51.8; p=0.69 | 33.5 vs. 13.9; p=0.33 | 0.9 vs. -6.7; p=0.443 | -4.5 vs. -17.2; p=0.54 |
|  | Unmethylated | 73.3 vs. 67.7; p=0.086 | 97.3 vs. 81.4; p=0.48 | 68.9 vs. 67.7; p=0.936 | 62.6 vs. 39.6; p=0.187 | 24 vs. 13.7; p=0.629 | -4.5 vs. 0.1; p=0.712 | -10.7 vs. -28.1; p=0.258 |
|  | Methylated+unmethylated | 69.2 vs. 68.3; p=0.749 | 97.9 vs. 82.1; p=0.234 | 67.4 vs. 65; p=0.756 | 61.6 vs. 45.7; p=0.196 | 28.8 vs. 13.8; p=0.242 | -1.8 vs. -3.3; p=0.827 | -7.6 vs. -22.6; p=0.19 |
| 34Gy | Methylated | 69.3 vs. 69.3; p=1.0 | 107.2 vs. 99; p=0.584 | 66.2 vs. 77.2; p=0.556 | 79.3 vs. 64.1; p=0.639 | 37.8 vs. 29.6; p=0.537 | -3.1 vs. 7.9; p=0.505 | 9.9 vs. -5.3; p=0.593 |
|  | Unmethylated | 70.3 vs. 67.7; p=0.294 | 115.4 vs. 83.3; p=0.078 | 76.7 vs. 53.8; p=0.021* | 68.9 vs. 41.1; p=0.02* | 45 vs. 15.6; p=0.073 | 6.4 vs. -13.9; p=0.023* | -1.4 vs. -26.6; p=0.036* |
|  | Methylated+unmethylated | 69.8 vs. 68.5; p=0.581 | 111.3 vs. 91.1; p=0.061 | 71.4 vs. 65.6; p=0.568 | 74.1 vs. 53.6; p=0.18 | 41.4 vs. 22.6; p=0.051 | 1.6 vs. -3; p=0.621 | 4.3 vs. -15.9; p=0.163 |
| 60Gy | Methylated | 60.3 vs.<br>71.7;<br>p=0.0003* | 107.7<br>vs. 106;<br>p=0.958 | 83.7 vs.<br>73.3;<br>p=0.442 | 82.8 vs.<br>55;<br>p=0.389 | 47.3 vs.<br>34.4;<br>p=0.671 | 23.4 vs.<br>1.6;<br>p=0.156 | 22.5 vs.<br>-16.7;<br>p=0.251 |
|  | Unmethylated | 66.7 vs.<br>68;<br>p=0.735 | 114.2<br>vs.<br>102.5;<br>p=0.649 | 78.2 vs.<br>63.1;<br>p=0.361 | 52 vs. 54;<br>p=0.931 | 47.6 vs.<br>34.5;<br>p=0.603 | 11.5 vs.<br>-4.9;<br>p=0.318 | -14.7 vs.<br>-14.1;<br>p=0.977 |
|  | Methylated+<br>unmethylated | 63.5 vs.<br>69.8;<br>p=0.023* | 111 vs.<br>104.3;<br>p=0.696 | 80.9 vs.<br>68.2;<br>p=0.183 | 67.4 vs.<br>54.5;<br>p=0.474 | 47.5 vs.<br>34.5;<br>p=0.444 | 17.4 vs.<br>-1.6;<br>p=0.061 | 3.9 vs.<br>-15.4;<br>p=0.307 |
| RT | Methylated | 64.8 vs.<br>70.5;<br>p=0.085 | 107.4<br>vs.<br>102.5;<br>p=0.735 | 74.9 vs.<br>75.3;<br>p=0.977 | 81 vs.<br>59.5;<br>p=0.277 | 42.6 vs. 32;<br>p=0.466 | 10.1 vs.<br>4.7;<br>p=0.627 | 16.2 vs.<br>-11;<br>p=0.161 |
|  | Unmethylated | 68.5 vs.<br>67.8;<br>p=0.756 | 114.8<br>vs. 93;<br>p=0.124 | 77.4 vs.<br>58.5;<br>p=0.028* | 60.5 vs.<br>47.5;<br>p=0.27 | 46.3 vs. 25;<br>p=0.119 | 8.9 vs.<br>-9.36;<br>p=0.031* | -8 vs. -20.3;<br>p=0.269 |
|  | Methylated+<br>unmethylated | 66.7 vs.<br>69.1;<br>p=0.192 | 111.1<br>vs.<br>97.7;<br>p=0.165 | 76.1 vs.<br>66.9;<br>p=0.169 | 70.8 vs.<br>53.5;<br>p=0.131 | 44.4 vs.<br>28.5;<br>p=0.091 | 9.5 vs. -2.3;<br>p=0.084 | 4.1 vs.<br>-15.7;<br>p=0.078 |

\*p<0.05

